# Assessing the impact of housing improvement and ivermectin administration to cattle on malaria transmission in southwest Ethiopia: A study protocol for a community-based cluster randomized control trial

**DOI:** 10.1101/2023.12.01.23299280

**Authors:** Fekadu Massebo, Betelihem Jima, Nigatu Eligo, Feven Wudneh, Mohammed Seid, Daniel Woldeyes, Biniam Wondale, Teklu Wegayehu, Bernt Lindtjørn

## Abstract

**Introduction:** The transmission of malaria and other infectious diseases can be linked to housing conditions. For malaria, poor housing conditions can result in higher indoor transmission rates compared to improved houses. The current study aims to introduce a new approach known as the push-pull strategy. This strategy involves screening houses to prevent mosquitoes from entering the houses and administering ivermectin to cattle to kill mosquitoes in the household compound. With this approach, we anticipate reducing mosquito bites and, subsequently, malaria incidence.

**Methods:** This study is a cluster randomized control trial in malaria-endemic *Kebeles* (villages –the smallest administrative unit) near the southern Rift Valley Lakes in the Gamo Zone of Ethiopia. The trial is open-labeled and four-armed with 60 clusters. The unit of randomization is a cluster (“*Limat Budin*” with 30-35 households) with an equal number of households in each arm. Each cluster will receive one of the following interventions: house screening (n=15), ivermectin cattle treatment (n=15), a combination of house screening and ivermectin cattle treatment (n=15), or no intervention (control arm, only a standard malaria prevention intervention; n=15). All clusters will continue with the essential malaria interventions such as bed nets and, according to the Ministry of Health policy. A total of 1912 households with 9060 individuals will be included in the study. The study’s primary outcome is determining malaria incidence among all age groups in the intervention groups compared to the control arm.

Over two years, we will screen study participants every four months to measure epidemiologic and serologic endpoints. Additionally, we will conduct bimonthly entomological assessments in 480 households with equal numbers in each arm for two years. All household members will undergo malaria testing using microscopy, with results confirmed through molecular methods to determine malaria prevalence and incidence. Children under five will also be tested for anemia with the Hemocue 301+ analyzer. We will use serological markers and entomological indices to estimate the human exposure to parasites and mosquito bites. Furthermore, we will evaluate the interventions durability, community acceptance, cost-effectiveness and it effect on household poverty reduction. We will perform an intention-to-treat analysis for cluster-level analysis.

**Discussion:** This is the first randomized control trial to assess the effectiveness of the push-pull strategy in controlling malaria in Ethiopia. This innovative approach aims to decrease the number of malaria-carrying mosquitoes indoors and outdoors, reduce human exposure to mosquito bites and parasites, and ultimately lower malaria incidence. Moreover, this strategy has the potential to ease the burden of malaria illness and aid in reducing household poverty.

**Ethics:** The trial has been approved by the Institutional Research Ethics Review Board (IRB/1423/2023) and the Animal Ethics Review Committee (AMU/AREC/12/2015) of Arba Minch University.

**Dissemination:** The study findings will be disseminated through presentations at national and international conferences, sharing information with study participants and stakeholders, and publications in peer-reviewed journals. Additionally, policymakers will be informed of the results and possibly incorporate them into the national malaria control toolbox.

**Trial registration:** The study is registered in the Pan African Clinical Trial Registry (PACTR202306667462566).

## Background and rationale

Malaria is a severe illness caused by *Plasmodium* parasites transmitted to humans through the bites of infected female *Anopheles* mosquitoes. In many countries with high malaria rates, using insecticide-treated nets (ITNs) and indoor residual spraying (IRS) has helped reduce the number of cases. However, in recent years, there has been an increase in cases [1]. Several factors may be contributing to this increase, including the impact of the COVID-19 pandemic on healthcare services and resources [1]. Additionally, many malaria-carrying mosquitoes have become resistant to various insecticides [2]. In areas with high coverage and the use of current interventions, some malaria mosquitoes resort to feeding on animals if humans are not available. These mosquitoes may bite humans during the early hours, leading to residual malaria transmission [3,4]. While interventions like ITNs and IRS target indoor biting and resting, outdoor biting and animal feeding can maintain the transmission [5]. Despite the combination of ITNs and IRS, malaria incidence has not decreased compared to using either intervention alone [6,7]. To eliminate malaria in Ethiopia and other affected countries, new strategies must be added to the control toolbox to strengthen elimination efforts [8].

Research efforts should focus on developing new control methods and enhancing sustainable development solutions, such as improving housing conditions [9]. Malaria and other vector-borne diseases disproportionately affect economically disadvantaged populations living in poorly constructed homes and unsatisfactorily managed environments. A recent systematic review on housing and vector-borne diseases indicates that housing improvement protects people against malaria and dengue infection [10]. Interestingly, improved housing protects everybody inside the house, can easily be integrated with existing interventions, reduces exposure to pollutants, and can improve indoor ventilation [11]. Our previous studies on housing intervention reduced the indoor density of vectors and the incidence of malaria [12,13]. Moreover, housing improvement prevents the entry of flies that carry enteric pathogens such as bacteria, viruses, protozoa, and fungi that can cause public health problems [13].

It is recommended to combine efforts to maximize benefits and target multiple diseases. To achieve this, we supplement the housing intervention, which diverts mosquitoes away from human dwellings, with interventions that attract mosquitoes. Ivermectin is a widely used treatment for endo- and ectoparasites in animals and for filarial nematode parasites in humans. Studies have demonstrated the effectiveness of ivermectin against malaria vectors [14,15], and its distinct mode of action sets it apart from commonly used insecticides for malaria control, making it a potential treatment option even for vectors that are resistant to frequently used insecticides like pyrethroids [16].

Studies have shown that improving housing can protect against diseases such as malaria, respiratory infections, and diarrhea [11,17,18]. Additionally, using ivermectin cattle treatment can reduce the number of mosquitoes that carry malaria [16,19,20]. However, there have yet to be any community trials combining these two malaria prevention interventions. While house screening has been proven to be cost-effective against malaria [21], it remains unclear whether combining it with administering ivermectin to cattle is effective. It is crucial to determine the additional expenses and cost-effectiveness of screening houses and treating cattle with ivermectin separately and together. Conducting these cost-effectiveness analyses is essential for policymaking and optimizing resource allocation in countries with limited accessibility [22].

We hypothesize that using these new push-pull interventions for malaria control can decrease the density of malaria-carrying mosquitoes indoors and outdoors, reduce human exposure to mosquito bites and parasites, and reduce malaria incidence. Moreover, it could reduce the malaria sickness burden and help multidimensional poverty reduction.

### Study objectives

#### Primary objective

The primary objective of this study is to determine whether house screening and ivermectin administration to cattle belonging to the household reduce malaria incidence among all age groups compared to the standard malaria control tools.

#### Secondary objectives

In the same study population, the secondary objectives of the trial are:

- To determine whether house screening and ivermectin cattle treatment reduce the density of host-seeking and resting malaria vectors.
- To determine whether house screening and ivermectin cattle treatment reduce malaria parasitemia among all age groups.
- To assess whether house screening and ivermectin cattle treatment reduce gametocyte carriage rates among all age groups.
- To assess the effect of house screening and ivermectin cattle treatment on the human blood index of malaria mosquitoes.
- To assess whether house screening and ivermectin cattle treatment reduce the gametocyte positivity rate in freshly fed mosquitoes.
- To assess the night-time mosquito biting and human activities in intervention and control arms.
- Using serological markers to determine whether house screening and ivermectin cattle treatment reduce human exposure to malaria mosquito bites.
- To assess the effect of house screening and ivermectin cattle treatment on malaria seroconversion rate among children under five years
- To assess the effect of house screening and ivermectin cattle treatment on human exposure to malaria parasites.
- To determine the effect of house screening and ivermectin cattle treatment on the spatiotemporal distribution of malaria and anemia.
- To assess the impact of house screening and ivermectin cattle treatment on bed net use rate.
- To assess the community acceptance of house screening and ivermectin cattle treatment malaria intervention.
- To determine the durability of the house screening intervention.
- To determine the cost-effectiveness of house screening and ivermectin cattle treatment against malaria compared to control arms.
- To evaluate the impact of house screening and ivermectin cattle treatment against malaria on the reduction of multidimensional poverty at the household level.
- To estimate household willingness to pay for the novel malaria intervention vis-a-vis cost of intervention.
- To assess whether house screening reduces the entry of other mosquito species and domestic flies.
- To investigate the effect of house screening on indoor temperature, carbon dioxide concentration, and relative humidity.

### Trial design

The design of this study is a cluster-randomized controlled trial with four arms. The cluster, which consists of approximately 30-35 households, will be randomly assigned into one of four arms in a 1:1:1:1 ratio (Fig 1). The design follows the SPRIT guideline (supporting information file S1). The clusters will be divided into intervention and control arms, with approximately equal number of households and individuals in each group. There will be a minimum of a 500 meters buffer zone between groups to prevent the spread of malaria vectors. All households and household members within each cluster will be included in the study to minimize selection bias and improve the generalizability [23]. Additionally, strict supervision will be in place to reduce observer bias.

**Fig 1.**
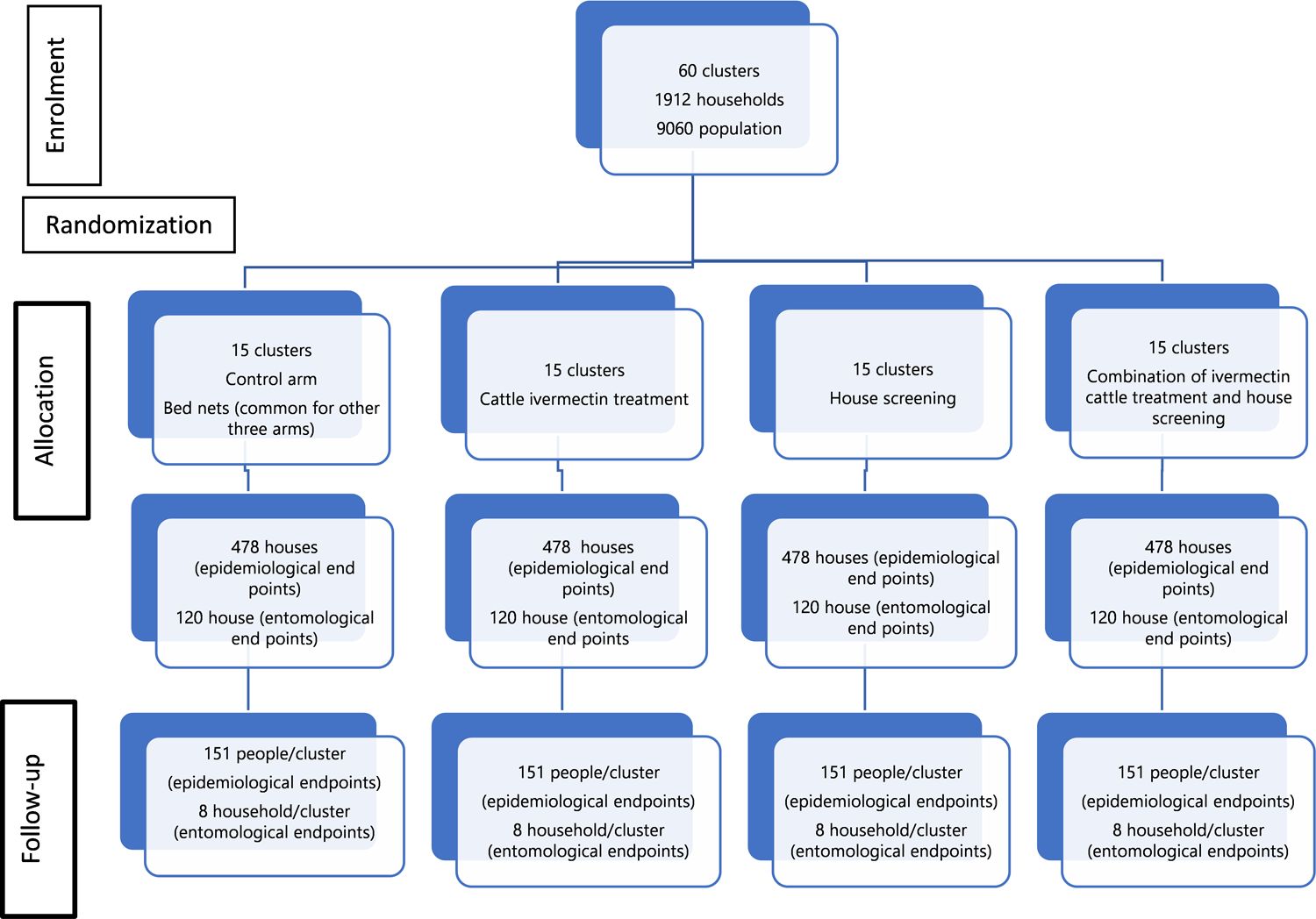
Study flowchart

The trial will have 60 clusters in four arms, with 15 clusters in each arm. One thousand nine hundred twelve households will be included for parasitological, serological, and cost-effectiveness analysis. An equal number of houses will be included in all arms to avoid imbalance. Maximum effort will be made to minimize the loss to follow-up in all arms. Four hundred eighty households will be used for entomological sampling, with an equal number of households in each arm.

## Methods: Participants, interventions, and outcomes

### Study setting

This study trial will be conducted in two districts of the Gamo Zone, located in southwest Ethiopia, where malaria is prevalent. Fifteen malaria-endemic rural and semi-urban *Kebeles* (villages - the smallest administrative unit) located close to the two southern Rift Valley Lakes, Lake Abaya and Lake Chamo, will be included (Fig 2). Most houses are constructed with wood, mud floors, mud walls, and corrugated iron roofs. Currently, grass-thatched traditional houses are rare and have been replaced by corrugated iron roof houses. The main source of income of the residents is agriculture, primarily banana, mango, maize, and tomato cultivation, often by irrigation from Abaya, Chamo Lakes, and tributary rivers such as Hare, Basso, Shefe, Elgo, and Sile.

**Fig 2.**
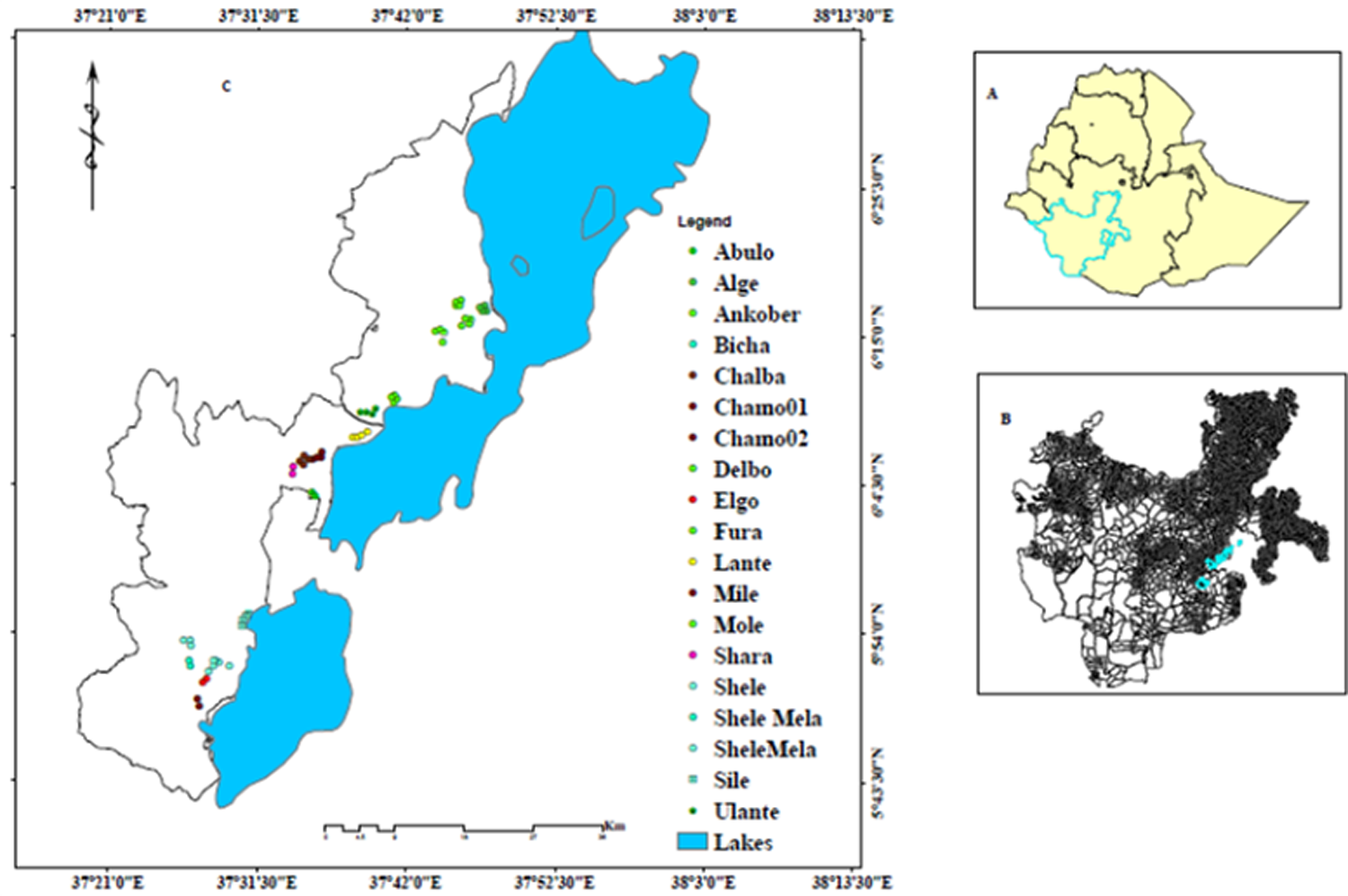
Location of the study villages (C), Southern Nations Nationalities and Peoples Region (B), Ethiopia (A).

Rainfall is bimodal, with the heaviest rains falling from April to May and the shorter rainy season occurring from September to December. Most malaria transmission occurs following rainfall, with two transmission peaks in a year. Though *P. falciparum* and *P. vivax* malaria parasites are occurring in the region, *P. falciparum* is the dominant parasite [24]. The primary malaria vector in the area is *An. arabiensis*, with *An. pharoensis* acting as a secondary vector [25–27]. According to the WHO bioassay test, *An. arabiensis* has developed resistance to pyrethroid insecticides [28,29]. Insecticide-treated bed nets (ITNs) and indoor residual spraying (IRS) are the two principal malaria control intervention tools deployed by the government [30]. The community gets access to primary health services from health posts in each village. These health posts have basic materials for immediate malaria diagnosis using rapid diagnostic tools (RDT). Anti-malarial drugs are available for the primary treatment of malaria cases.

### Eligibility criteria

All households and their members will be randomly assigned to either the intervention or control groups. Participants who provide informed consent will be included in the two-year malaria study. To minimize variation between villages, the study will focus on villages situated near the shores of the two Rift Valley lakes that are endemic for malaria. Mosquitoes’ flight distance will also be considered when selecting clusters to investigate to reduce the diversion effect of the intervention and contamination resulting from mosquito movement [31].

Any households that decline to participate in the study or have housing structures not conducive to screening will be excluded. Additionally, we will not include any cattle that are lactating or planned to be used for meat shortly. Calves under six months of age, and pregnant cows will also be excluded from the study.

### Interventions

All houses in the intervention clusters will receive either house screening, cattle treatment, or a combination of house screening and cattle treatment. All arms will be provided with ITNs. The number of bed nets in each house will be proportional to the number of household members by considering one-bed net for two individuals [1]. Those who receive only bed nets will be used as a control group.

In the house screening arm, 15 clusters will receive house screening intervention to block the entry of malaria vectors. Following discussions with communities and household heads, metal wire mesh will be fixed permanently on windows externally. Ventilation holes will be covered by wire mesh, with good size for good airflow. Wooden-framed doors with wire mesh will be permanently installed on the main door frame and will open outward. The wire mesh will receive a coat of anti-rust paint to prevent corrosion damage. Homeowners will be reminded to keep their screened doors closed at night through regular follow-up. Trained artisans from each study village will oversee the screening process. House residents will receive training on caring for the screens and avoiding activities that could damage them. Monthly inspections will be conducted to determine the screens’ integrity. Study participants will be encouraged to keep the screened doors closed all the time.

In the clusters designated for the endectocide drug ivermectin, all cattle will receive injectable ivermectin (0.2 mg/kg) every two months. Veterinarians working in the villages will administer the treatment. Ivermectin is a licensed drug for treating endoparasites and ectoparasites in livestock, which improves animal health and productivity. It also has a different mode of action than insecticides used for malaria vector control, reducing the risk of cross-resistance [16]. Grazing in the same field will not cause contamination as systemic endectocide will be used. Treating cattle for malaria control is recommended as the principal malaria vector, *An. arabiensis* tends to feed on cattle [32]. In the study region, high livestock density could maximize the benefits of this livestock-based intervention.

The use of insecticide-treated nets (the current best practice in malaria control) will be consistent across all groups and will serve as the comparator for comparing the effectiveness of cattle treatment, house screening, and a combination of both. In addition, the effectiveness of the combination of cattle treatment and house screening will be compared to the individual interventions of cattle treatment and house screening.

### Outcomes

#### Primary outcome variables

- Incidence of malaria in all age groups
- Prevalence of malaria in all age groups

#### Secondary outcome variables

##### Epidemiology and social

- Prevalence of gametocyte carriage in human hosts
- Prevalence of anemia in children aged between 6 months and 5 years
- Spatiotemporal distribution of malaria and anemia
- Community acceptance of the intervention
- The durability of house screening
- ITNs use rate

##### Entomology and serological

- Indoor and outdoor density of malaria mosquitoes
- Prevalence of antibodies against salivary gland antigens
- Seroprevalence of malaria parasites
- Entomological inoculation rate
- Sporozoite infection rate
- Blood meal index
- Mosquito biting rates
- The proportion of freshly fed mosquitoes with gametocyte
- Effectiveness of serological biomarkers in monitoring vector control
- Human night activities and sleeping patterns (time)
- Indoor density of other mosquitoes and flies

##### Economic evaluation

- Cost-effectiveness of the intervention
- Household multidimensional poverty reduction
- Willingness to pay for the intervention

##### Other outcomes

- Prevalence of animal ectoparasites

##### Adverse effect outcome variables

- Prevalence of respiratory infection rate
- Indoor room temperature
- Indoor air carbon dioxide concentration
- Indoor air relative humidity

### Participant timeline

Figure 3 displays the timetable for enrollment, interventions, and evaluations.

**Fig 3.**
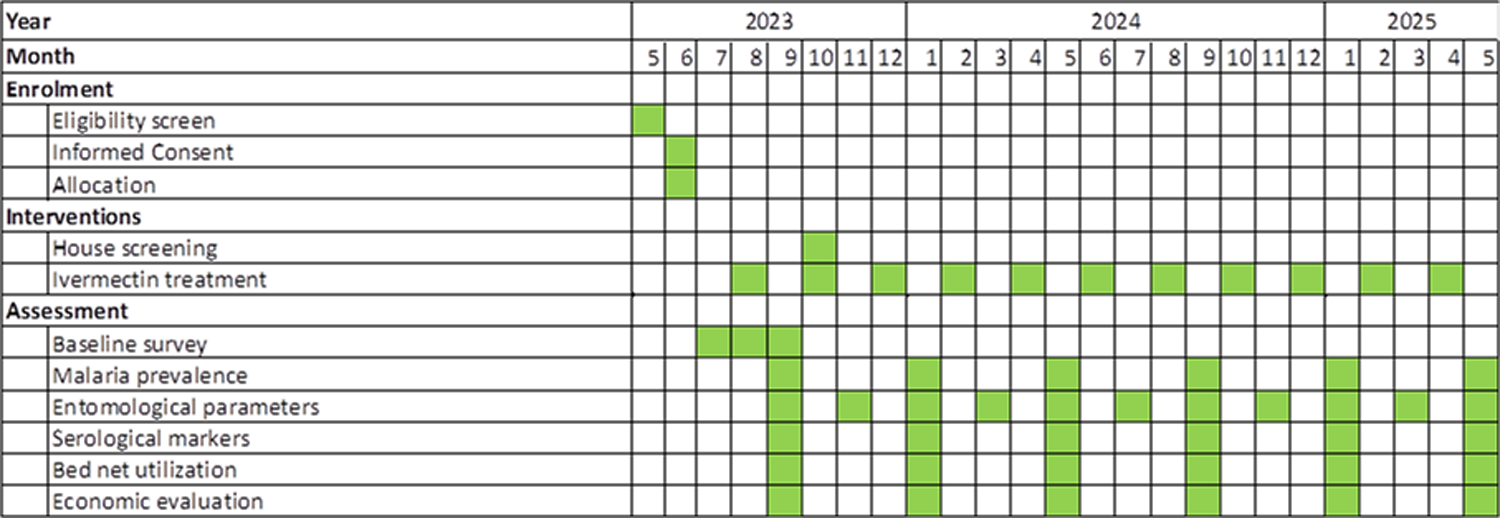
The trial timetable for enrollment, interventions, and evaluations.

### Sample size

#### Epidemiological study

The sample size will be determined following the assumptions by Hayes and Bennet [33] for incidence rate. The 50% reduction of incidence rate, the equal sample size in all arms, 80% power, 0.05 significant levels, and 0.5 coefficient of variation will be assumed to determine the sample size. Using the above information, we estimated to include 478 houses in each arm (Figure 1). We will include 1912 houses (on average 32 houses per cluster) in the trial, with an estimated 9060 participants (151 individuals in each cluster) across 60 clusters.

#### Entomological study

According to a study in the area, indoor hosting-seeking mosquitoes were reduced by 48% due to screening doors and windows [12]. Our estimation suggests that a combination of house screening and treatment of cattle with systemic insecticide could have an even greater impact, leading to a 60% reduction in malaria vectors. To conduct an entomological study with a 5% significant level, 90% power, and an equal number of houses in each group, we will include 480 houses - 120 houses in each arm.

#### Recruitment

After obtaining legal permission from respective administrative offices, a community sensitization and awareness creation meeting will be performed. The objective of the study will be explained to staff at each level. Each household head will be informed about the house screening and ivermectin cattle treatment’s aim, benefits, and risks. All consented household members will be invited and recruited to participate in the study.

After obtaining written consent, the household will be given a unique identification (ID) number, which will be tagged onto a colored metal plate and placed on the upper front door of the house. Each inhabitant in the household will be informed to use this ID number whenever they need assistance from the project.

## Methods: assignment of interventions

### Allocation

The study will randomize 60 clusters into four arms, with 15 clusters in each group. Each cluster will include 30-35 households. To ensure balance, block randomization will be used to allocate interventions to four clusters in each village, with all intervention types being used in each village. Balancing each group will help to reduce imbalances due to the heterogeneity of malaria transmission. The clusters will be randomized using SPSS software and a computer-generated seed number by personnel outside the study area. One group will receive house screening and cattle treatment, another group will receive only cattle treatment, a third group will receive only house screening, and the fourth group will receive bed nets, a common intervention in all groups. To prevent contamination and minimize the impact of mosquito movement, there will be a buffer zone of at least 500m between clusters. All individuals in the selected communities will be advised to use bed nets.

### Blinding/Masking

Due to the nature of the intervention, study participants and investigators will not be blinded. The laboratory staff and those involved in data analysis will be blinded to minimize performance bias. Strict supervision will minimize observer bias, and continuous follow-up will encourage the house owners to keep their doors closed at night and engage in cattle treatment.

## Methods: data collection, management, analysis

### Data collection methods

Data collection tools will be developed and arranged in a way that can address research questions. At the start of the study, a baseline census of the population will be conducted. Each house found in a cluster will be mapped by spatial coordinates. A unique ID number will be provided for each household and individual. After participant enrolment and intervention in all clusters are correctly applied, actual data collection for assessing the impact of interventions will be performed. Structured questionnaires translated into the Amharic language will be used for demographic and clinical data collection. The questionnaire will be pretested in the neighboring district before data collection. During the pre-test, frequent review meetings will be conducted with data collectors and supervisors to assess the clarity and completeness of the questions; then, the results will be documented. The actual data collection process will be strictly supervised. The supervisors will monitor the activities of each data collection process concurrently with data collectors to increase trustworthiness.

The study employed locally trained data collectors who ensure cultural sensitivity and understanding. They will visit each designated home to gather the required data. If the participants are unavailable during the initial visit, two more visits will be scheduled to reduce the loss of follow-up. The Kobo Toolbox will be used to conduct the data collection process. The research team and data manager can only access the data server.

### Epidemiological and serological data collection

All household members will be screened by microscopy for malaria three times a year for two years. These repeated visits will enable us to collect malaria data in all transmission seasons and improve the generalizability of the study. Asymptomatic and symptomatic cases will be recorded. Every positive case will receive immediate treatment at their respective village health post. The axillary temperature will be measured and recorded.

The collected blood sample will be used for microscopic slide preparation for parasite detection and quantification, field measurement of anemia, and Dried Blood Spot (DBS) preparation in Cellulose Chromatography filter paper (Fishery Scientific) for further molecular and immunological testing and storage in RNA solution for gametocyte detection. Each DBS sample will be labeled and allowed to air dry overnight at room temperature. The sample will be individually placed in zip-lock plastic bags (25 cm × 25 cm) with silica desiccant gel (Sigma) and transported to Arba Minch University Medical Entomology and Vector Control Molecular Laboratory.

A thick blood smear will be prepared for malaria parasite detection, while a thin smear will be for species identification. The blood films will be stained in the nearby health centers with an experienced laboratory technologist, who will not participate in slide reading. Two independent expert laboratory technologists will read the slides for parasite identification and *Plasmodium* parasite density quantification. The microscopists will be blinded to the participants’ information. Any discrepancy will be resolved by PCR test from DBS. PCR will screen microscopy-negative cases for final confirmation. Parasite density of symptomatic and asymptomatic cases will be quantified. RTqPCR will confirm the gametocyte carriage rate of symptomatic and asymptomatic patients.

The DBS sample will undergo an indirect enzyme-linked immunosorbent assay (ELISA) antibody test to evaluate for exposure to the malaria parasite. Biomarkers such as apical membrane antigen 1 (AMA-1), merozoite surface protein antigens (MSP-119), and Etramp5 will be used for this purpose [34,35]. As a proxy measure of human exposure to malaria mosquito bites, we will measure the IgG response specific to the salivary gland antigens (gSG6-P1) from *An. gambiae* mosquitoes [36]. In the field, the hemoglobin (Hb) concentration will be measured directly using capillary blood samples with a Hemocue®Hb 301+ analyser (HemoCue AB, Angelholm, SE).

### Entomological data collection

Entomological sampling will be conducted every other month using CDC light traps, Pyrethrum Spray Catches (PCS), pit shelters, and Human Landing Catches (HLC) in randomly selected households across all arms for two years. To ensure impartiality due to human skills, CDC light trap collection techniques will be used to assess the effectiveness of the intervention.

Indoor and outdoor host-seeking mosquito collection will be conducted using CDC light traps [23]. To assess the density of mosquitoes resting inside, we will use PSC, while pit shelters will be used for those resting outside. Mosquitoes will be identified under a microscope using a morphological key [37]. Our laboratory staff will screen the mosquitoes for circum-sporozoite protein (CSPs) species identification and to determine the origin of their blood meal. We will also determine the biting patterns of malaria mosquitoes by collecting them through HLC. We will use CDC light traps to estimate the density of host-seeking malaria mosquitoes indoors and outdoors.

We will conduct direct observations using questionnaires to study human night-time activity and sleeping patterns. We will select 20% of households participating in the CDC light trap for in-depth interviews. We will also measure HLC in adjacent clusters to determine if the intervention impacts the density of host-seeking malaria mosquitoes in nearby areas not included in the study. The study will consist of houses on the intervention’s outskirts and nearby clusters not part of the trial.

### Health economic data collection and analysis

Financial records from the project finance will be used to collect cost-related data. The healthcare sector perspective will be taken into account, including household’s out-of-pocket costs. In addition, a limited societal perspective will also be considered to account for cost components beyond those captured by the healthcare sector perspective. These components may include transportation, unpaid caregiver time, and productivity loss. To estimate the per capita costs of the intervention, we will divide the average cost per house by the household size. This will be compared to the per capita costs of routine practice. We will estimate each household’s Quality Adjusted Life Years (QALY) based on the standard methods, using the number of malaria cases averted [38]. The incremental cost-effectiveness ratio (ICER) will be calculated by dividing the difference in the per capita cost between the intervention and control groups by the difference in their QALY gained. In order to assess how a novel intervention against malaria affects household poverty, we will utilize the Multidimensional Poverty Index (MPI). This method is useful for identifying which specific aspects of poverty, or combinations of aspects, have been reduced, rather than solely focusing on single factor. To assess the sustainability of the novel malaria intervention, the study will use the willingness-to-pay approach of households concerning the intervention cost.

### Data management and access to data

Arba Minch University will take overall responsibility for the project’s ongoing development, review, and implementation of the Data and Digital Outputs Management Plan. The Data Management Coordinating Centre will oversee the intra-study data-sharing process. All researchers in the project will be given access to the cleaned data sets. All data collection will be done through the Kobo Toolbox system (https://support.kobotoolbox.org/kobocollect-android.html).

All data sets will be password-protected. Each researcher will have direct access to their own data sets and access to other data by request. After the project ends, we shall share the data, ensuring that the data safeguarding policy is not breached in sharing and storing data. During the project’s life cycle (before migrating data to an open access repository), data will be held by Arba Minch University’s storage area network, comprised of servers in physically secure data centers, following institutional policy on data safeguarding.

To share data and maximize its accessibility, data will be made publicly available through relevant data repositories. After the study ends, study protocols, all publications, and data will be archived and made publicly available, and each of these items will have a Digital Object Identifier [DOI]. In collaboration with the Data Centre, efforts will be made to ensure that data is provided and organized in a format that is accessible. Only anonymized data will be made publicly available. In the case of household survey data, we will follow standard protocols for anonymization.

### Statistical methods

We will use descriptive statistics such as frequency and percentage to describe the population characteristics. For cluster-level analysis of outcome variables, we will perform an intention-to-treat analysis. To compare outcome variables between intervention and control clusters, we will use a multilevel mixed-effect logistic regression model and consider the clustering effect. We will assess the crude and adjusted odds ratio with their corresponding 95% CI and a P-value of <0.05 to control the confounding factors, clustering effect of villages, and measurement effects. To account for the dispersion of mosquitoes, we will apply a Generalized Estimating Equation (GEE) with a negative binomial error distribution. In addition, we will use a first-order autoregressive correlation structure to account for a serial correlation between repeated catches made in the same house. To determine the percentage reduction of house entry, we will calculate the mean ratio of mosquitoes between intervention and control houses. Following Consolidated Standards of Reporting Trials (CONSORT) guidance, we will report trial outcome data[39].

## Ethics and dissemination

### Research ethic approval

The Institutional Research Ethics Review Board (IRB/1423/2023) and the Animal Ethics Review Committee (AMU/AREC/12/2015) of Arba Minch University have approved the study trial protocol. Additionally, permission letters were acquired from various administrative offices of the health department in Gamo Zone. Prior to enrollment into the study, participants were fully informed about the study objectives and procedures, and written informed consent was obtained. The risks and benefits of the study will be informed to all household heads and members. Any study participants testing positive for malaria will be treated immediately in the nearby health post according to National guidelines [40].The protocol has been registered at the Pan African Clinical Trials port (PACTR202306667462566).

### Protocol amendments

Once the protocol has been accepted, any changes can only be made with the agreement of the principal investigator, sponsor and the IRB. Amendments will only be done if absolutely necessary and every effort will be made to minimize any bias resulting from the changes.

### Consent/assent

Before starting this trial, permissions will be obtained from the zonal, district, and village administrators. The objectives of the study will be explained to all concerned bodies. Written and oral consent will be sought before performing any procedure to enrolment into the study participants. For participants under the age of 18, a consent form was also signed by their parent, legal guardian, or person with power of attorney. The principal investigator ensured that the participants and their legally acceptable representative comprehended the information and answered any questions about the study. Participants were notified that giving consent was purely voluntary and could not be influenced by any form of coercion.

### Confidentiality

All study participants’ data will be kept confidential before, during, and after the study by encoding the participants. Data dispersed to project team members will be blinded to identifying participant information to ensure confidentiality.

### Ancillary and post-trial care

In the event of a severe adverse effect during the trial, the principal investigator will be promptly informed and the participant will receive immediate care according to the reporting procedures. As per the Declaration of Helsinki [39], at the end of the study, households in the control group will have the choice to screen their households. Additionally, households in the intervention group may opt to have the screens removed from their homes if they desire.

### Criteria for discontinuing or modifying allocated interventions

The household head can withdraw cattle for slaughter or milk whenever necessary. Additionally, if screening doors and windows restricts airflow and leads to respiratory issues and other medical problems are revealed, they are advised to withdraw them immediately. Moreover, if the participants want to return their house to the pre-intervention state, it will be returned to the previous state.

### Relevant concomitant care permitted or prohibited during the trial

The purpose of this study is to assess how effective a combination of house screening and cattle treatment is in reducing malaria transmission. To ensure accurate results, the use of partial IRS application will be prohibited. In the control clusters, the treatment of cattle and screening of houses will also be prohibited. Additionally, intentional damage to the screened doors and windows will be strictly prohibited.

### Oversight and monitoring

#### Risks

The benefits and risks of screening houses and treating cattle will be explained. The indoor carbon dioxide level and heat will be monitored if there is variation due to house screening intervention. An interim analysis will not be conducted, but the data management team will keep an eye on the impact of the interventions and any reported side effects. It is believed that these interventions pose minimal risks as they have been proven and widely implemented [18,40,41].

#### Strategies to improve adherence to the interventions

Loss to follow-up occurs when an enrolled participant will not attend the scheduled visits. Maximum effort will be made to minimize the loss to follow-up. Community sensitization and mobilization will be done to ensure the maximum participation of the study participants. The benefits they get from cattle treatment will be explained repeatedly through community meetings. Continuous supervision will be done to ensure the study participants’ adherence.

#### Withdrawal of consent

A participant may withdraw consent at any time. If someone does not follow the intervention instructions or experiences severe adverse effects, they can withdraw. In this case, information on the adverse event and symptomatic treatment will be recorded in a case report form. If the adverse event is severe, the principal investigator will be notified immediately and follow the reporting procedures described in the guidelines. The reasons for discontinuation or protocol violation will be recorded on the case report form.

#### Discontinuation or protocol violation

The investigator might withdraw a participant from the study intervention and follow-up procedures if the individual or household in the cluster violated the protocol.

### Monitoring and evaluation

The study plan shall be monitored regularly to assess the coverage of interventions and their impact and determine whether programmes are proceeding as intended or adjustments are required. The committee will maintain a record of any negative effects observed in the health centers. This will enable them to keep track of the health outcomes of the programme and promptly identify the need for corrective actions if required. Annual, mid-term, and final reviews will be conducted to evaluate project progress with stakeholders.

### Quality assurance

The following will be considered: Training for data collection and pilot study, supervision and pre-testing of tools, and laboratory quality control.

#### Avoidance of bias

The study will include all residents within each cluster to avoid any bias in selection. To account for the differences between study villages, clusters within each village will be randomly assigned in a block. We will make every effort to ensure that participants stay engaged throughout the study to minimize dropouts. To maintain objectivity, external study assistants will blind the documentation of the primary endpoint, and the success of blinding will be verified. The study protocol will be made public.

### Information flow and dissemination communication

Internal communication will be facilitated through bi-monthly meetings and a monitoring system. Communications across the project team will be through a project mailing list, and the team will share project documents, work plans, and results. The team will summarize activities and project highlights based on the monitoring and reporting system before each meeting.

Researchers and others affiliated with the project will submit abstracts to relevant national and international conferences during the project to share results with the broader community. The project will support Ethiopian researchers to participate in conferences. The findings will be presented in locally organized workshops, and the project’s major findings will be distributed to the district where the project can be implemented. The results will also be published in peer-reviewed Journals.

## Discussion

ITNs and IRS helped decrease malaria cases and deaths until 2015 [42], but the number of cases remained stable until 2019 [43]. Despite this, malaria is still prevalent, and residual transmission may occur indoors and outdoors [44]. Studies have shown that improving housing conditions can help reduce the density of malaria vectors and protect all household members [11, 18, 43]. This can be combined with interventions such as bed nets and indoor spraying to control residual malaria.

Research has demonstrated that using wire mesh on doors, windows, and ventilation holes can reduce the density of *An. arabiensis* mosquitoes indoors [13]. A recent study in the area found that screening doors and windows reduced the indoor density of *An. arabiensis*. In the intervention houses, the entomological inoculation rate of *An. arabiensis* and malaria incidence was lower than in control houses [12]. The intervention cost per individual was 6.5 USD, and most participants in the intervention arm were willing to continue using screened doors and windows.

The effectiveness of housing interventions can be maximized by supplementing them with a mosquito-pulling strategy. This includes treating cattle with ivermectin to kill mosquitoes feed on animals [15,16]. Ivermectin is a highly effective insecticide against malaria mosquitoes that is resistant to other treatments [46,47]. Ivermectin is a safe and effective solution for combatting malaria suitable for vertebrates, including humans and cattle. What sets it apart from other insecticides is that it does not affect the chloride-gated iron channels group, making it a safe option for animals. Recent research indicates that incorporating ivermectin-based interventions into vector control strategies can significantly reduce both the prevalence and incidence of malaria [14,15]. In Ethiopia, where the primary malaria vector feeds on cattle [32], utilizing ivermectin treatment for cattle can be an invaluable tool for curbing the spread of this disease.

This research examines how effectively the push-pull strategy reduces malaria incidence and the density of malaria-carrying mosquitoes indoors and outdoors. Additionally, the study will assess how this approach affects the exposure of humans to mosquito bites and malaria parasites using serological markers. The findings of this investigation will be helpful for policymakers both nationally and internationally to expand the malaria vector control toolbox.

## Data Availability

After the study ends, study protocols, all publications, and data will be archived and made publicly available, and each of these items will have a Digital Object Identifier [DOI]. In collaboration with the Data Centre, efforts will be made to ensure that data is provided and organized in a format that is accessible. Only anonymized data will be made publicly available. In the case of household survey data, we will follow standard protocols for anonymization

## Acknowledgments

We thank the Gamo Zone Health Department and the Arba Minch Zuria and Mirab Abaya districts Health Offices. We also extend our appreciation to the Birbir, Lante, and Kolla Shelle Health Centers for their assistance in mobilizing the community for discussion and their commitment to supporting the implementation of the intervention.

## Authors’ contributions

Conceptualization: Fekadu Massebo, Bernt Lindtjørn

Funding acquisition: Fekadu Massebo, Bernt Lindtjørn

Methodology: Fekadu Massebo, Bernt Lindtjørn

Project administration: Fekadu Massebo, Bernt Lindtjørn

Supervision: Fekadu Massebo, Teklu Wegayehu, Biniam Wondale, Daniel Woldeyes, Bernt Lindtjørn

Writing: Fekadu Massebo, Betelihem Jima, Nigatu Eligo, Feven Wudneh, Mohammed Seid, Biniam Wondale, Daniel Woldeyes, Teklu Wegayehu, Bernt Lindtjørn

Writing – review & editing: Fekadu Massebo, Betelihem Jima, Nigatu Eligo, Feven Wudneh, Mohammed Seid, Daniel Woldeyes, Biniam Wondale, Teklu Wegayehu, Bernt Lindtjørn

## Trial status

The collection of baseline data is currently in progress.

